# Long-term follow-up of treatment comparisons in RECOVERY: a randomised, open-label, platform trial for patients hospitalised with COVID-19

**DOI:** 10.1101/2025.08.29.25334732

**Authors:** Peter W Horby, Leon Peto, Mark Campbell, Rachel Wade, Guilherme Pessoa-Amorim, Vanessa Tobert, Natalie Staplin, Jonathan R Emberson, Karl Wallendszus, William M Stevens, Andy King, Rijo Kurien, Charles Crichton, Christopher Brightling, Benjamin Prudon, Christopher A Green, Sheri Thackoorcharan, Paul Hine, Tim Felton, Richard Stewart, Heinke Kunst, Andrew Ustianowski, J Kenneth Baillie, Maya H Buch, Saul N Faust, Thomas Jaki, Katie Jeffery, Edmund Juszczak, Marian Knight, Wei Shen Lim, Alan Montgomery, Aparna Mukherjee, Andrew Mumford, Kathryn Rowan, Guy Thwaites, Marion Mafham, Richard Haynes, Martin J Landray, RECOVERY Collaborative Group

## Abstract

**Background:** The Randomised Evaluation of COVID-19 Therapy (RECOVERY) trial evaluated the effects of sixteen potential treatments for patients hospitalised with COVID-19. Dexamethasone (at a dose of 6mg daily), tocilizumab, baricitinib, and the monoclonal antibodies casirivimab-imdevimab and sotrovimab were shown to reduce 28-day mortality in all or specific groups of patients. Here we report the long-term efficacy and safety of all sixteen therapies.

**Methods:** Patients hospitalised with COVID-19 were potentially eligible to join this randomised, controlled, open-label, platform trial. Participants were randomly allocated to receive each trial treatment, or not, on top of usual care. Analyses were by intention to treat comparing each treatment with its own usual care control group. The pre-specified primary long-term follow-up outcome was 6-month all-cause mortality, presented as mortality rate ratios adjusted for baseline age and ventilation status. The key safety outcomes were major non-COVID infection and non-COVID death at 6 months. ISRCTN50189673 and NCT04381936.

**Findings:** Between 19 March 2020 and 19 March 2024, 48,402 patients were included in RECOVERY COVID-19 treatment comparisons. For each of the treatments previously demonstrated to be effective at 28 days, the early mortality benefit was preserved up to 6 months. Among 6425 patients in the dexamethasone (6mg daily) comparison, 6-month mortality was 34.3% vs 44.4% in the invasive mechanical ventilation group (rate ratio [RR] 0.68; 95% confidence interval [CI] 0.55–0.85; p=0.0006); 27.7% vs 29.2% in the oxygen or non-invasive ventilation group (RR 0.87; 95% CI 0.77–0.99; p=0.034); and 26.1% vs 22.5% in the no oxygen group (RR 1.10; 95% CI 0.89–1.36; p=0.39); test for trend p=0.0024. Among 4116 patients in the tocilizumab comparison, 34.3% vs 38.9% died within 6 months (RR 0.87; 95% CI 0.79–0.96; p=0.0077). Among 8156 patients in the baricitinib comparison, 15.7% vs. 16.6% died (RR 0.89; 95% CI 0.80–0.99; p=0.032). Among 3153 anti-SARS-CoV-2 serum antibody negative patients (the primary analysis population) in the casirivimab-imdevimab comparison, 29.3% vs. 34.7% died (RR 0.87; 95% CI 0.77–0.98; p=0.024). Among 720 patients with high serum nucleocapsid antigen concentration (the primary analysis population) in the sotrovimab comparison, 33.0% vs. 38.6% died (RR 0.78; 95% CI 0.61–1.00; p=0.050). In line with the 28-day results, aspirin, azithromycin, colchicine, convalescent plasma, dimethyl fumarate, empagliflozin, lopinavir-ritonavir, molnupiravir, and nirmatrelvir-ritonavir did not reduce 6-month mortality. Mortality at 6 months was higher with hydroxychloroquine therapy (33.3% vs. 30.2%; RR 1.14; 95% CI 1.02–1.27; p=0.018) and, among hypoxic patients not requiring ventilatory support, with higher dose dexamethasone (initial dose 20mg daily, 22.0% vs. 17.6%, RR 1.34; 95% CI 1.04–1.72; p=0.021). Allocation to dexamethasone 6mg once daily resulted in a small increase in major non-COVID infection within 6 months compared with usual care (21.4% vs. 19.1%; absolute difference 2.2%; 95% CI 0.2–4.5%). We found no evidence that any other treatments increased the risk of major non-COVID infection.

**Interpretation:** In patients hospitalised with COVID-19, dexamethasone (at a dose of 6mg daily in hypoxic patients), tocilizumab (in hypoxic patients with CRP ≥75 mg/L), baricitinib, casirivimab-imdevimab (in seronegative patients), and sotrovimab (in high antigen patients) reduced 6-month mortality. Dexamethasone at a dose of 6mg daily was associated with an increase in major non-COVID infection but there was no evidence of other later emerging harms. Other treatments tested in RECOVERY did not reduce 6-month mortality.

**Funding:** UK Research and Innovation (Medical Research Council) and National Institute for Health and Care Research (Grant ref: MC_PC_19056), and Wellcome Trust (Grant Ref: 222406/Z/20/Z).

## INTRODUCTION

The RECOVERY trial was initiated early in the COVID-19 pandemic as a means to generate reliable evidence on the effects of potential treatments on 28-day mortality for patients hospitalised with COVID-19. The trial has now evaluated sixteen treatments, five of which were shown to reduce 28-day mortality.^1–5^ Three of the effective treatments were immunomodulators: the corticosteroid dexamethasone (at a dose of 6mg once daily, among hypoxic patients), the interleukin-6 (IL-6) antagonist tocilizumab (among hypoxic patients with CRP ≥75 mg/L), and the Janus kinase (JAK) inhibitor baricitinib. The other two effective treatments were antiviral monoclonal antibodies: casirivimab-imdevimab (among individuals without detectable antibodies to the SARS-CoV-2 spike protein), and sotrovimab (among patients with high serum SARS-CoV-2 nucleocapsid antigen levels). Dexamethasone, tocilizumab and baricitinib are currently used in clinical practice to treat severe COVID-19.^6,7^ Use of casirivimab-imdevimab and sotrovimab has been limited by their loss of activity against subsequent SARS-CoV-2 variants.

The remaining evaluated treatments – aspirin, azithromycin, colchicine, convalescent plasma, dimethyl fumarate, empagliflozin, higher dose dexamethasone (at an initial dose of 20mg once daily among hypoxic patients needing ventilatory support), hydroxychloroquine, lopinavir-ritonavir, molnupiravir, and nirmatrelvir-ritonavir did not demonstrate clinical benefits at 28-day follow-up.^8–17^ Among hypoxic patients who were not receiving ventilatory support at randomisation, higher dose dexamethasone was associated with an increased 28-day mortality, which led to the early closure of the comparison for these patients.^18^

Long-term follow-up is important to ascertain whether the demonstrated benefits of treatments at 28 days are sustained over time and to assess whether any evaluated treatments are associated with later hazards, in particular secondary infection. Secondary infection complicating COVID-19 is a common and serious complication, developing in about one quarter of hospitalised patients with COVID-19 and is associated with higher mortality.^19,20^ Observational studies have postulated an association between secondary infections and COVID-19 immunomodulatory treatments, but residual confounding by indication is likely, emphasising the need for randomised data.^20,21^ The risk of complications following COVID-19 hospitalisation remains high even after discharge, with around 10% patients being readmitted, and 7% dying within 90 days.^22^

Here, we report the results of a randomised evaluation of sixteen COVID-19 therapies on 6-month mortality (the pre-specified primary long-term outcome) and on non-COVID infections and non-COVID causes of death (the key safety objectives).

## METHODS

### Study design and participants

The RECOVERY trial is an investigator-initiated, individually randomised, controlled, open-label, platform trial that evaluated treatments for patients hospitalised with COVID-19 between 19 March 2020 and 31 March 2024 (when funding for COVID-19 treatment evaluations ended).^23^ The RECOVERY platform continues to evaluate treatments for influenza and community-acquired pneumonia. Details of each COVID-19 treatment comparison, including the primary 28-day outcomes, have been published previously.^1–5,8–18^ The COVID-19 comparisons took place at 175 hospital organisations in the United Kingdom, supported by the National Institute for Health and Care Research Clinical Research Network, as well as at 23 hospitals in South and Southeast Asia and Africa (appendix pp2-30). The trial is coordinated by the Nuffield Department of Population Health at the University of Oxford (Oxford, UK), the trial sponsor. The trial is conducted in accordance with the principles of the International Conference on Harmonisation–Good Clinical Practice guidelines. The protocol was approved by all relevant regulatory authorities and ethics committees in each participating country (appendix p31). The current statistical analysis plan and first and final versions of the protocol are available in the appendix (pp91-204) with additional information, including all previous versions of the protocol and statistical analysis plan, available on the study website (www.recoverytrial.net/results) and previous publications.^1–5,8–18^

Patients admitted to hospital were eligible for the study if they had clinically suspected or laboratory confirmed SARS-CoV-2 infection (laboratory confirmation was required from 29 November 2021), and no medical history that might, in the opinion of the attending clinician, put the patient at significant risk if they were to participate in the trial. Further inclusion and exclusion criteria for each individual treatment comparison are shown in the appendix (pp32-33). Children recruited to evaluations of treatments for Paediatric Inflammatory Multisystem Syndrome Temporarily associated with SARS-CoV-2 are not included in these analyses, with results published separately.^24^ Written informed consent was obtained from all patients, or a legal representative if patients were too unwell or otherwise unable to provide informed consent.

### Randomisation and masking

Eligible and consenting patients were assigned to either usual standard of care plus the relevant active treatment or usual standard of care alone, using web-based simple (unstratified) randomisation with allocation concealed until after randomisation. The dosing regimens of active treatments are shown in the appendix (p32-33).

As a platform trial with factorial randomisation, patients could be simultaneously included in assessments of other treatments that they were eligible for and that were open at the time (appendix pp43-45). Participants and local study staff were not blinded to the allocated treatment. Other than members of the Data Monitoring Committee, all individuals involved in the trial were blinded to aggregated outcome data while recruitment and 28-day follow-up were ongoing and were blinded to subsequent aggregated outcome data until data collection was completed for 6-month follow-up analyses. The statistical analysis plan for the long-term follow-up outcomes was finalised prior to seeing relevant unblinded results.

### Procedures

Baseline data were collected using a web-based case report form that included demographics, level of respiratory support, major comorbidities, suitability of the study treatment for a particular patient, and treatment availability at the study site (appendix p48).

For the initial follow-up period, a single online follow-up form was completed when participants were discharged, had died or at 28 days after randomisation, whichever occurred earliest (appendix p50). Information was recorded on adherence to allocated study treatment, receipt of other COVID-19 treatments, duration of admission, receipt of respiratory or renal support, and vital status (including cause of death). In addition, in the UK, routine healthcare and registry data were obtained including information on vital status (with date and cause of death), discharge from hospital, receipt of respiratory support, and renal replacement therapy (appendix pp205-229). For sites outside the UK, a further case report form (appendix p62) collected vital status at day 28.

For long-term follow-up (beyond 28 days), routine healthcare and registry data were used in the UK, additionally collecting information on diagnoses associated with the initial hospital stay or readmission, and blood culture and respiratory tract microbiology results. The final death certification, hospital coding and microbiology datasets used for the analysis were received in March 2024 (99.8% of participants were randomised >6 months before this). For sites outside the UK, a 6-month follow-up case report form collected equivalent information, including vital status and details of causes of extension of hospital stay or readmission (appendix p64).

Linkage to routine healthcare data was used to identify patients who were severely immunocompromised at trial entry. Definitions followed UK criteria for SARS-CoV-2 vaccination and shielding for people highly vulnerable due to immunodeficiency, including patients with haematological malignancy, solid organ or haematological transplant, hyposplenism, and those on immunosuppressive medication (appendix p34).

### Outcomes

For these long-term follow-up analyses the pre-specified primary outcome was all-cause mortality at 6 months, and the pre-specified key safety outcomes were major non-COVID infection and non-COVID causes of death, both at 6 months.

Major non-COVID infection was defined by any of the following: (i) non-COVID infection reported on a follow-up case report form; (ii) non-COVID infection recorded as a primary cause of death on the death certificate; (iii) non-COVID infection code from post-randomisation hospital care (index admission or readmission) indicating a potentially serious infection (excluding codes for chronic viral hepatitis and uncomplicated superficial skin, upper respiratory or gastrointestinal infections); or (iv) clinically important blood or respiratory tract culture from a post-randomisation sample. Where possible, infections were categorised according to (i) anatomical site and (ii) the type of infecting organism (bacterial, fungal, viral or other). Further details are in appendix pp220-221. Underlying cause of death was obtained from the UK Office for National Statistics (appendix pp216-217).

Pre-specified exploratory analyses were performed at 6 months for the secondary and subsidiary outcomes reported at 28 days in the primary trial publications. Secondary outcomes were: (i) time to discharge alive from hospital, and (ii) new requirement for invasive mechanical ventilation or death (among patients not on invasive mechanical ventilation at randomisation). Subsidiary outcomes were: (i) new requirement for non-invasive respiratory support (among patients not on ventilatory support at randomisation), (ii) time to successful cessation of invasive mechanical ventilation (among those on invasive ventilation at randomisation), and (iii) new requirement for renal dialysis or haemofiltration (among patients not receiving renal replacement therapy at randomisation). A pre-specified exploratory analysis was performed of post-randomisation hospital diagnoses recorded as the primary reason for a period of in-patient care (during the index admission or a readmission, categorised by ICD-10 chapter). Further details about the derivation of hospital diagnoses are available in appendix p220.

### Statistical analysis

The primary analysis for all outcomes was by intention to treat, comparing patients allocated trial treatment with those not allocated trial treatment, but for whom that treatment was suitable and available (i.e. concurrent controls). For the 6-month all-cause mortality and non-COVID mortality outcomes, the hazard ratio from a Cox model adjusted for age and level of respiratory support was used to estimate the mortality rate ratio (RR), with sensitivity analyses for the primary outcome of all-cause mortality conducted with additional adjustment for other key baseline pre-specified subgroups. We also constructed Kaplan-Meier survival curves adjusted for age and level of respiratory support to display cumulative mortality over the long-term follow-up period, which was extended to 24 months (appendix p35). Participants outside the UK were censored on day 184 as no further follow-up data was available beyond 6 months. We used the same method to analyse time to hospital discharge and successful cessation of invasive mechanical ventilation, with patients who died in hospital right-censored on day 184. For the composite outcome of progression to invasive mechanical ventilation or death within 6 months (among those not receiving invasive mechanical ventilation at randomisation), and the outcomes of receipt of ventilation and use of haemodialysis or haemofiltration, the precise dates were not available and so a log-binomial regression model was used to estimate the age and respiratory support adjusted risk ratio. Estimates of rate and risk ratios are shown with 95% confidence intervals.

For the analyses of major non-COVID infection, cause-specific mortality, and hospital recorded diagnoses, counts and percentages by randomised group are presented overall and by pre-specified sub-categories. Adjusted absolute risk differences are presented in which the age and respiratory support adjusted RR (or its lower or upper 95% CI) is applied to the absolute risk in the usual care arm.

For the casirivimab-imdevimab comparison, the prespecified primary analysis population was participants without detectable serum antibodies to the SARS-CoV-2 spike protein (seronegative) at randomisation. The prespecified primary analysis population for the sotrovimab comparison was participants with serum SARS-CoV-2 nucleocapsid antigen concentrations greater than the median at randomisation.

A limited number of prespecified subgroup analyses were performed for the primary outcome with an appropriate statistical test for interaction (heterogeneity or trend), defined by the following characteristics at randomisation: level of respiratory support (for the dexamethasone comparisons), concomitant use of corticosteroids (for the tocilizumab and baricitinib comparisons), concomitant use of IL-6 antagonists (for the baricitinib comparison), recipient anti-SARS-CoV-2 spike protein antibody status (for the casirivimab-imdevimab comparison), and SARS-CoV-2 antigen level (for the sotrovimab comparison). In addition, we performed exploratory subgroup analyses of the effects study treatments on mortality and major non-COVID infection by baseline immunodeficiency status.

All p-values are two-sided and are shown without adjustment for multiple testing. The full database is held by the study team which collected the data from study sites and performed the analyses at the Nuffield Department of Population Health, University of Oxford (Oxford, UK).

Analyses were performed using SAS version 9.4 and R version 4.2.3. The trial is registered with ISRCTN (50189673) and clinicaltrials.gov (NCT04381936).

### Role of the funding source

The funders of the study had no role in study design, data collection, data analysis, data interpretation, or writing of the report. The corresponding authors had full access to all the data in the study and had final responsibility for the decision to submit for publication.

## RESULTS

A total of 48,402 patients were recruited across sixteen COVID-19 treatment comparisons in RECOVERY between 19 March 2020 and 19 March 2024 (Figure 1, appendix p87). Baseline characteristics have been described previously and are summarised in Table 1 and appendix pp68-69. The mean age of participants across the whole trial was 62.8 years (SD 15.6) (Table 1). 97% were recruited in the UK, 3% in Asia and <0.5% in Africa. Corticosteroid treatment became a component of usual care in June 2020 and was received by around 90% of patients randomised after this. The declining incidence of severe COVID-19 in the UK from early 2022 onwards meant that later treatment comparisons were smaller than those conducted earlier in the pandemic, with more uncertainty around treatment effect estimates (appendix p87). After 6 months of follow-up, 46,302 (96%) participants had known vital status based on their available sources of data. Across all treatment comparisons combined, a total of 10,132 (21%) participants died within 28 days and a further 2,063 (4%) died within 6 months.

**Figure 1:**
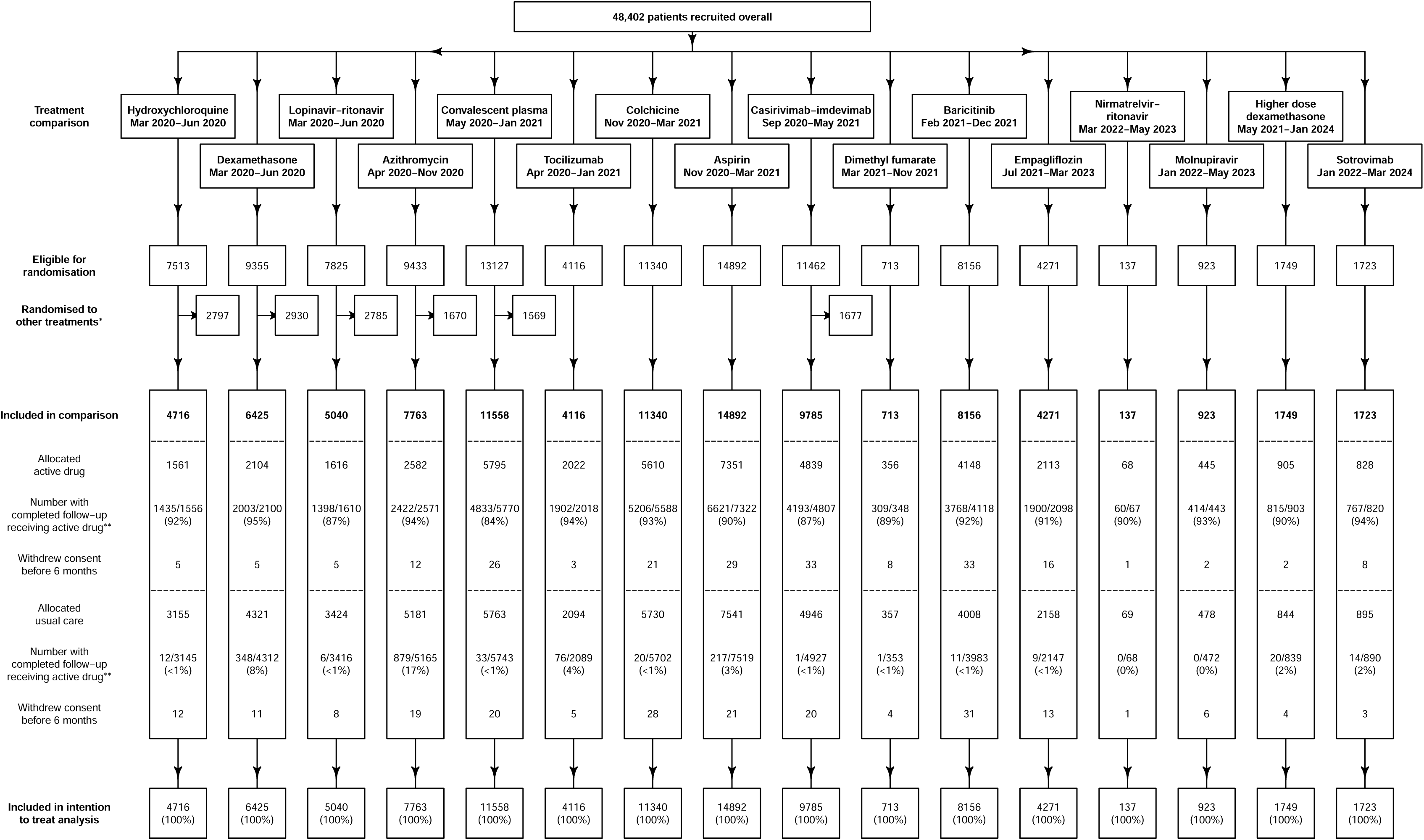
Trial profile. When multiple treatment comparisons were suitable and available, participants could enter more than one comparison (see appendix pp43-45 for the randomisation scheme throughout the trial). * Patients allocated an alternative active treatment in randomisations that included two or more active treatments as well as usual care. Does not include patients allocated to other treatment in factorial (independent) randomisations. ** Denominator is participants with a completed follow-up form. For the dexamethasone comparison, active drug includes any corticosteroid. For the azithromycin comparison it includes any macrolide. For the tocilizumab comparison it includes tocilizumab or sarilumab. For the higher dose dexamethasone comparison, it includes patients who received dexamethasone at a dose of 20mg or higher (760/839 [91%] participants allocated usual care in that comparison received lower doses of corticosteroids, mostly 6mg dexamethasone daily).

**Table 1:** Baseline characteristics for randomised comparisons involving treatments shown to reduce 28-day mortality.

|  | Treatment comparison |  |  |  |  |  |
| --- | --- | --- | --- | --- | --- | --- |
|  | Dexamethasone<br>(n=6425) | Tocilizumab<br>(n=4116) | Baricitinib<br>(n=8156) | Casirivimab-Imdevimab<br>(Seronegative)<br>(n=3153) | Sotrovimab<br>(High antigen)<br>(n=720) | Overall*<br>(n=48402) |
| First patient enrolled in comparison | Mar-2020 | Apr-2020 | Feb-2021 | Oct-2020 | Jan-2022 | Mar-2020 |
| Last patient enrolled in comparison | Jun-2020 | Jan-2021 | Dec-2021 | May-2021 | Jan-2024 | Mar-2024 |
| Age, years | 66.1 (15.7) | 63.6 (13.6) | 58.1 (15.5) | 63.6 (15.4) | 72.3 (13.5) | 62.8 (15.6) |
| <70** | 3646 (57%) | 2685 (65%) | 6228 (76%) | 1997 (63%) | 264 (37%) | 31673 (65%) |
| ≥70 to <80 | 1329 (21%) | 959 (23%) | 1319 (16%) | 692 (22%) | 244 (34%) | 9798 (20%) |
| ≥80 | 1450 (23%) | 472 (11%) | 609 (7%) | 464 (15%) | 212 (29%) | 6931 (14%) |
| Sex |  |  |  |  |  |  |
| Male | 4090 (64%) | 2775 (67%) | 5378 (66%) | 1873 (59%) | 444 (62%) | 30454 (63%) |
| Female † | 2335 (36%) | 1341 (33%) | 2778 (34%) | 1280 (41%) | 276 (38%) | 17948 (37%) |
| Region and ethnicity |  |  |  |  |  |  |
| UK | 6425 (100%) | 4116 (100%) | 8156 (100%) | 3153 (100%) | 720 (100%) | 46835 (97%) |
| White | 4823 (75%) | 3171 (77%) | 6625 (81%) | 2621 (83%) | 636 (88%) | 36620 (78%) |
| BAME †† | 1151 (18%) | 757 (18%) | 953 (12%) | 295 (9%) | 48 (7%) | 7102 (15%) |
| Unknown | 451 (7%) | 188 (5%) | 578 (7%) | 237 (8%) | 36 (5%) | 3113 (7%) |
| Asia ‡ | 0 | 0 | 0 | 0 | 0 | 1515 (3%) |
| Africa § | 0 | 0 | 0 | 0 | 0 | 52 (<0.5%) |
| Number of days since symptom onset | 9 (5-13) | 10 (7-14) | 9 (6-11) | 7 (5-9) | 6 (3-12) | 9 (5-12) |
| Number of days since hospitalization | 2 (1-5) | 2 (1-4) | 1 (1-3) | 1 (1-2) | 2 (1-4) | 2 (1-3) |
| Respiratory support received at randomisation |  |  |  |  |  |  |
| No oxygen | 1535 (24%) | 9 (<0.5%) | 465 (6%) | 330 (10%) | 97 (13%) | 6002 (12%) |
| Simple oxygen or non-invasive ventilation ¶ | 3883 (60%) | - | - | - | - | 7956 (16%) |
| Simple oxygen | - | 1859 (45%) | 5513 (68%) | 2080 (66%) | 439 (61%) | 21621 (45%) |
| Non-invasive ventilation \|\| | - | 1686 (41%) | 1927 (24%) | 673 (21%) | 158 (22%) | 9568 (20%) |
| Invasive mechanical ventilation *** | 1007 (16%) | 562 (14%) | 251 (3%) | 70 (2%) | 26 (4%) | 3255 (7%) |
| Severely immunocompromised §§ |  |  |  |  |  |  |
| Yes | 436 (7%) | 257 (6%) | 409 (5%) | 301 (10%) | 262 (36%) | 3393 (7%) |
| No | 5834 (91%) | 3802 (92%) | 7370 (90%) | 2743 (87%) | 438 (61%) | 41799 (86%) |
| Unknown | 155 (2%) | 57 (1%) | 377 (5%) | 109 (3%) | 20 (3%) | 3210 (7%) |
| SARS-CoV-2 test result ¶¶ |  |  |  |  |  |  |
| Positive | 5750 (89%) | 3948 (96%) | 7858 (96%) | 3077 (98%) | 720 (100%) | 45860 (95%) |
| Negative | 647 (10%) | 141 (3%) | 73 (1%) | 34 (1%) | 0 | 1865 (4%) |
| Unknown | 28 (<0.5%) | 27 (1%) | 225 (3%) | 42 (1%) | 0 | 677 (1%) |
| Use of other treatments ‡‡ |  |  |  |  |  |  |
| Corticosteroids | - | 3385 (82%) | 7771 (95%) | 2883 (91%) | 662 (92%) | 36030 (74%) |
| Remdesivir | - | 1117 (27%) | 1667 (20%) | 765 (24%) | 272 (38%) | 9770 (20%) |
| Tocilizumab or sarilumab | - | - | 1872 (23%) | 110 (3%) | 126 (18%) | 2936 (6%) |

Results are count (%), mean ± standard deviation, or median (inter-quartile range).
\* Includes participants in all RECOVERY COVID-19 comparisons (including those shown in webtable 1)
\*\* Includes 99 patients aged <18 across all comparisons (these patients were excluded from the tocilizumab, colchicine, aspirin, high-dose dexamethasone, and empagliflozin comparisons). See specific reports for the number in each comparison.
† Includes 101 pregnant women across all comparisons (these patients were excluded from the colchicine, baricitinib, and empagliflozin comparisons). See specific reports for the number in each comparison.
†† UK black and minority ethnic (ethnicity not recorded at non-UK sites).
‡ Nepal, Indonesia, Vietnam, and India.
§ South Africa and Ghana.
¶ Category only used for patients recruited to non-tocilizumab comparisons before 21 September 2020 (before this the randomisation form made no distinction between simple oxygen and non-invasive ventilation).
|| Includes patients receiving high-flow nasal oxygen, continuous positive airway pressure, or other non-invasive ventilation.

For dexamethasone, mortality at 6 months was lower in patients allocated dexamethasone (6mg daily) versus usual care (596/2104 [28.3%] vs 1297/4321 [30.0%]; RR 0.87; 95% CI 0.79–0.96; p=0.0042; Figure 2, Table 2). There was significant heterogeneity in the proportional effect of dexamethasone according to the prespecified subgroups of baseline respiratory support (test for trend p=0.0024; appendix p88), similar to that previously reported at 28 days. Dexamethasone reduced the risk of death among patients receiving invasive mechanical ventilation (34.3% vs. 44.4%; RR 0.68; 95% CI 0.55–0.85; p=0.0006) and among those receiving oxygen or non-invasive ventilation (27.7% vs. 29.2%; RR 0.87; 95% CI 0.77–0.99; p=0.034), but not among those who did not require oxygen at randomisation (26.1% vs 22.5%; RR 1.10; 95% CI 0.89–1.36; p=0.39; Figure 2, Table 2, appendix p88). In an exploratory analysis, there was no evidence of heterogeneity of the effect of dexamethasone by baseline immunodeficiency status (appendix p89). There was an excess of major non-COVID infection in those allocated dexamethasone compared with those allocated usual care (451/2104 [21.4%] vs. 827/4321 [19.1%]; absolute difference 2.2%; 95% CI 0.2 to 4.5%; Table 2), with no evidence that this risk differed by level of respiratory support. No significant difference was observed in non-COVID mortality at 6 months (119/2104 [5.7%] vs 211/4321 [4.9%]; RR 1.08; 95% CI 0.86–1.35; Table 2).

**Figure 2:**
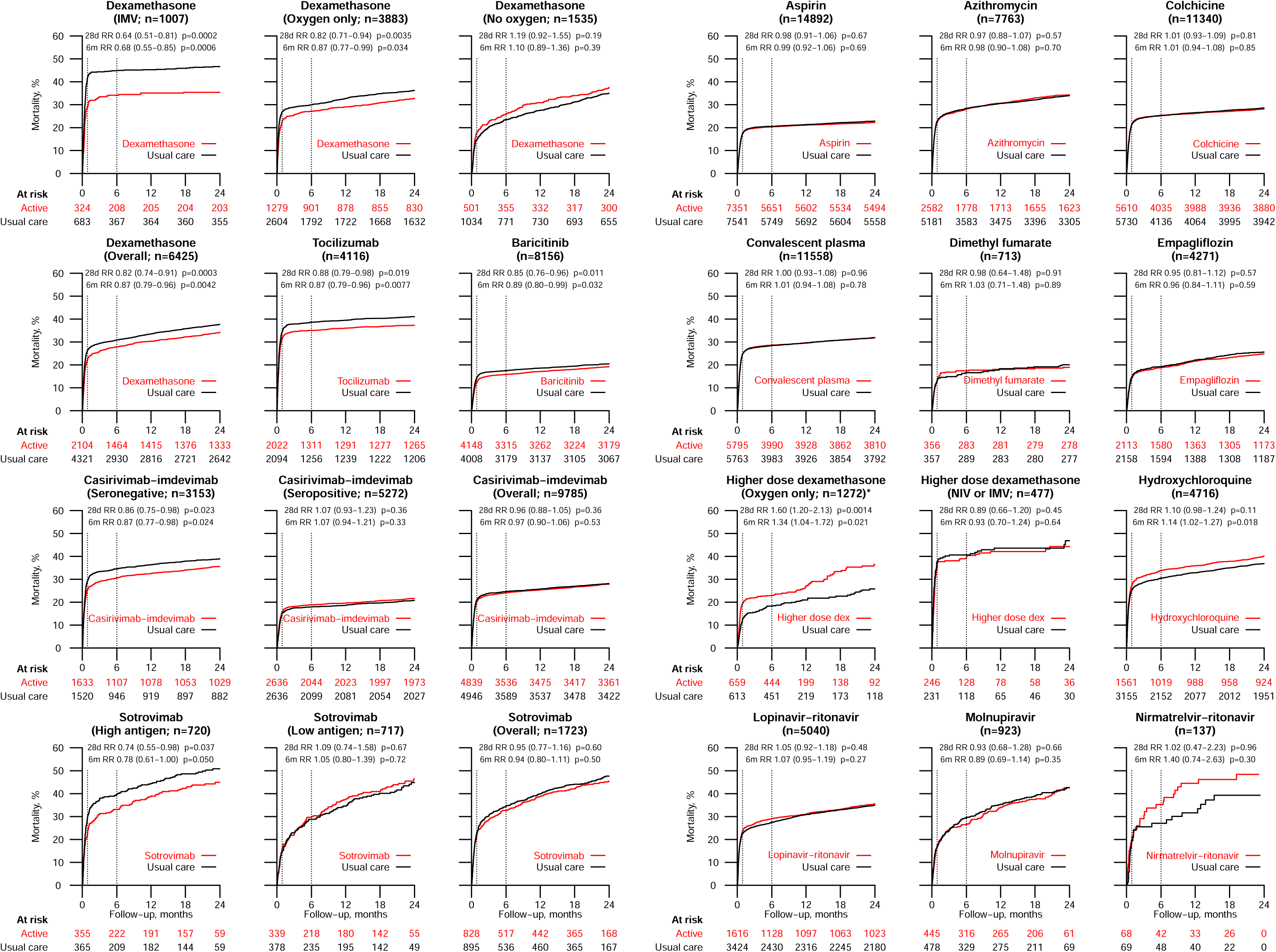
Kaplan-Meier survival curves for mortality, separately by treatment comparison and, if appropriate, in subgroups of interest. Dotted vertical lines are shown at 28 days and 6 months. Rate ratios (RRs) and p-values are adjusted for age (in three categories: <70 years, 70 to 79 years, and ≥80 years) and, if appropriate, level of respiratory support. IMV=Invasive mechanical ventilation. NIV=Non-invasive ventilation. *In the higher dose dexamethasone comparison, recruitment of patients not requiring ventilatory support was stopped early because of an apparent hazard in this subgroup, identified during a routine review of unblinded data by the Data Monitoring Committee, and results should be interpreted in the context of this data-dependent stop.

**Table 2:** Effect of treatments shown to reduce 28-day mortality on key 6-month efficacy and safety outcomes.

| Efficacy and safety outcomes at 6 months |  |  |  |  |  |  |  |  |  |  |
| --- | --- | --- | --- | --- | --- | --- | --- | --- | --- | --- |
| Treatment comparison |  | All-cause mortality |  |  | All major non-COVID infection |  |  | Non-COVID mortality |  |  |
|  |  | Active | Control | RR (95% CI) | Active | Control | RD (95% CI) | Active | Control | RR (95% CI) |
| Dexamethasone | IMV | 111/324 (34.3%) | 303/683 (44.4%) | 0.68 (0.55-0.85) | 118/324 (36.4%) | 231/683 (33.8%) | 2.8% (-3.2 to 9.8) | 5/324 (1.5%) | 8/683 (1.2%) | 1.11 (0.36-3.38) |
|  | Oxygen only | 354/1279 (27.7%) | 761/2604 (29.2%) | 0.87 (0.77-0.99) | 210/1279 (16.4%) | 385/2604 (14.8%) | 1.5% (-0.8 to 4.2) | 62/1279 (4.8%) | 108/2604 (4.1%) | 1.08 (0.79-1.47) |
|  | No oxygen | 131/501 (26.1%) | 233/1034 (22.5%) | 1.10 (0.89-1.36) | 123/501 (24.6%) | 211/1034 (20.4%) | 3.8% (-0.5 to 9.0) | 52/501 (10.4%) | 95/1034 (9.2%) | 1.08 (0.77-1.52) |
|  | Overall | 596/2104 (28.3%) | 1297/4321 (30.0%) | 0.87 (0.79-0.96) | 451/2104 (21.4%) | 827/4321 (19.1%) | 2.2% (0.2 to 4.5) | 119/2104 (5.7%) | 211/4321 (4.9%) | 1.08 (0.86-1.35) |
| Tocilizumab | Overall | 694/2022 (34.3%) | 815/2094 (38.9%) | 0.87 (0.79-0.96) | 377/2022 (18.6%) | 416/2094 (19.9%) | -0.8% (-3.0 to 1.7) | 44/2022 (2.2%) | 48/2094 (2.3%) | 0.91 (0.61-1.37) |
| Baricitinib | Overall | 650/4148 (15.7%) | 664/4008 (16.6%) | 0.89 (0.80-0.99) | 839/4148 (20.2%) | 764/4008 (19.1%) | 0.2% (-1.3 to 1.8) | 89/4148 (2.1%) | 77/4008 (1.9%) | 1.03 (0.76-1.40) |
| Casirivimab-imdevimab | Seronegative | 478/1633 (29.3%) | 527/1520 (34.7%) | 0.87 (0.77-0.98) | 358/1633 (21.9%) | 365/1520 (24.0%) | -0.8% (-3.4 to 2.2) | 68/1633 (4.2%) | 59/1520 (3.9%) | 1.03 (0.72-1.46) |
|  | Seropositive | 488/2636 (18.5%) | 457/2636 (17.3%) | 1.07 (0.94-1.21) | 464/2636 (17.6%) | 427/2636 (16.2%) | 1.4% (-0.5 to 3.5) | 46/2636 (1.7%) | 47/2636 (1.8%) | 0.99 (0.66-1.48) |
|  | Overall | 1124/4839 (23.2%) | 1203/4946 (24.3%) | 0.97 (0.90-1.06) | 950/4839 (19.6%) | 943/4946 (19.1%) | 0.9% (-0.5 to 2.5) | 130/4839 (2.7%) | 124/4946 (2.5%) | 1.07 (0.83-1.37) |
| Sotrovimab | High antigen | 117/355 (33.0%) | 141/365 (38.6%) | 0.78 (0.61-1.00) | 151/355 (42.5%) | 151/365 (41.4%) | 0.9% (-5.7 to 8.8) | 39/355 (11.0%) | 49/365 (13.4%) | 0.72 (0.47-1.10) |
|  | Low antigen | 95/339 (28.0%) | 105/378 (27.8%) | 1.05 (0.80-1.39) | 135/339 (39.8%) | 150/378 (39.7%) | -0.1% (-6.6 to 7.7) | 60/339 (17.7%) | 72/378 (19.0%) | 0.98 (0.69-1.38) |
|  | Overall | 263/828 (31.8%) | 300/895 (33.5%) | 0.94 (0.80-1.11) | 335/828 (40.5%) | 366/895 (40.9%) | -0.7% (-5.1 to 4.1) | 113/828 (13.6%) | 135/895 (15.1%) | 0.89 (0.69-1.15) |

RR=age- and respiratory support-adjusted rate ratio for all-cause and non-COVID mortality, RD=age- and respiratory support-adjusted risk difference for the outcome of any major non-COVID infection. Age adjusted for in three categories (<70 years, 70-79 years, and 80 years or older). CI=confidence interval.

For tocilizumab, mortality at 6 months was lower in those allocated tocilizumab compared with usual care alone (694/2022 [34.3%] vs. 815/2094 [38.9%]; RR 0.87; 95% CI 0.79–0.96; p=0.0077; Figure 2, Table 2). There was no significant heterogeneity by use of corticosteroids at randomisation (appendix p88). In an exploratory analysis, there was evidence of heterogeneity by baseline immunodeficiency status, with possible hazard in immunocompromised patients (heterogeneity p=0.0011, appendix p89). The occurrence of major non-COVID infection was similar in those allocated tocilizumab versus usual care (377/2022 [18.6%] vs. 416/2094 [19.9%]; absolute difference −0.8%; 95% CI −3.0 to 1.7%; Table 2). In an exploratory subgroup analysis, there was no evidence of heterogeneity by baseline corticosteroid use, or baseline immunodeficiency status (appendix p70, p90). No significant difference was observed in non-COVID mortality at 6 months (44/2022 [2.2%] vs 48/2094 [2.3%]; RR 0.91; 95% CI 0.61–1.37; Table 2).

For baricitinib, mortality at 6 months was lower in those allocated baricitinib compared with usual care alone (650/4148 [15.7%] vs. 664/4008 [16.6%]; RR 0.89; 95% CI 0.80– 0.99; p=0.032; Figure 2, Table 2). There was no evidence of heterogeneity by use of corticosteroids or IL-6 antagonists at randomisation (appendix p88). The occurrence of major non-COVID infection was similar in those allocated baricitinib versus usual care (839/4148 [20.2%] vs. 764/4008 [19.1%]; absolute difference 0.2%; 95% CI −1.3 to 1.8%; Table 2). In an exploratory subgroup analysis, there was no evidence of heterogeneity by baseline corticosteroid use, IL-6 use, or baseline immunodeficiency status (appendix p70-71, p90). No significant difference was observed in non-COVID mortality at 6 months (89/4148 [2.1%] vs 77/4008 [1.9%]; RR 1.03; 95% CI 0.76–1.40; Table 2).

For casirivimab-imdevimab, in the pre-specified primary analysis population of seronegative participants, mortality within 6 months was lower in the casirivimab-imdevimab group compared with the usual care group (478/1633 [29.3%] vs 527/1520 [34.7%]; RR 0.87; 95% CI 0.77–0.98; p=0.024; Figure 2, Table 2). In the overall population (regardless of serostatus), mortality was similar in the casirivimab-imdevimab group and usual care groups (1124/4839 [23.2%] vs 1203/4946 [24.3%]; RR 0.97; 95% CI 0.90–1.06; p=0.53). In an exploratory analysis, there was evidence of heterogeneity by baseline immunodeficiency status, with greater benefit in immunocompromised patients (heterogeneity p=0.011, appendix p89). Among the overall population, the occurrence of major non-COVID infection was similar in those allocated casirivimab-imdevimab versus usual care (950/4839 [19.6%] vs. 943/4946 [19.1%]; absolute difference 0.9%; 95% CI −0.5 to 2.5%; Table 2). No significant difference was observed in non-COVID mortality at 6 months (130/4839 [2.7%] vs 124/4946 [2.5%]; RR 1.07; 95% CI 0.83–1.37; Table 2).

For sotrovimab, in the pre-specified primary analysis population of participants with serum SARS-CoV-2 antigen levels above the median at randomisation, mortality within 6 months was lower in the sotrovimab group compared with the usual care group (117/355 [33.0%] vs 141/365 [38.6%]; RR 0.78; 95% CI 0.61–1.00; p=0.050; Figure 2, Table 2). In the overall population (regardless of antigen level), mortality was similar in the sotrovimab and usual care groups (263/828 [31.8%] vs 300/895 [33.5%]; RR 0.95; 95% CI 0.80–1.11; p=0.50). In an exploratory analysis, there was evidence of heterogeneity by baseline immunodeficiency status, with greater benefit in immunocompromised patients (heterogeneity p=0.0081, appendix p89). Among the overall population, the occurrence of major non-COVID infection was similar in those allocated sotrovimab versus usual care (335/828 [40.5%] vs. 366/895 [40.9%]; absolute difference −0.7%; 95% CI −5.1 to 4.1%; Table 2). No significant difference was observed in non-COVID mortality at 6 months (113/828 [13.6%] vs 135/895 [15.1%]; RR 0.89; 95% CI 0.69–1.15; Table 2).

Long-term follow-up analyses were also conducted for the treatment comparisons of aspirin, azithromycin, colchicine, convalescent plasma, dimethyl fumarate, empagliflozin, higher dose dexamethasone, hydroxychloroquine, lopinavir-ritonavir, molnupiravir, and nirmatrelvir-ritonavir. As was the case at 28 days, none of these treatments resulted in a significant 6-month mortality benefit compared with usual care (Figure 2). The increased 28-day mortality previously observed for higher dose dexamethasone in non-ventilated patients persisted at 6 months (145/659 [22.0%] vs 108/613 [17.6%]; RR 1.34; 95% CI 1.04 – 1.72; p=0.019; Figure 2). The evidence to suggest an increased mortality with hydroxychloroquine at day 28 was somewhat stronger at 6 months (520/1561 [33.3%] vs 953/3155 [30.2%]; RR 1.14; 95% CI 1.02–1.27; p=0.018; Figure 2), with a numerical excess seen in most causes of COVID and non-COVID deaths (appendix p77). Otherwise there was no evidence that any of these other treatments affected the risk of non-COVID death. The rate of major non-COVID infection was lower in patients receiving azithromycin (412/2582 [16.0%] vs 932/5181 [18.0%]; absolute difference −2.1%; 95% CI −3.6 to −0.3%), mostly related to a lower rate of bacterial infection (appendix p72). Otherwise, there was no evidence that any of these other treatments affected the risk of major non-COVID infection.

Across all comparisons, no safety concerns were identified when comparing post-randomisation hospital diagnoses in patients allocated active treatment versus usual care (appendix pp78-81). The effects of all trial treatments on secondary and subsidiary outcomes at 6 months were similar to those seen previously in 28-day analyses (appendix pp82-85). A sensitivity analysis of 6-month mortality that included adjustment for all key baseline pre-specified subgroups produced near-identical results to the main analyses (data not shown).

## DISCUSSION

Longer-term follow-up of RECOVERY participants indicates that the early mortality benefits observed for three immunomodulatory treatments (dexamethasone, tocilizumab, baricitinib) and two monoclonal anti-viral treatments (casirivimab-imdevimab and sotrovimab) were maintained over 6 months. Dexamethasone (6mg daily) was associated with a small increase in the rate of major non-COVID infection, of around 2%, but this should be viewed in the context of a clear reduction in all-cause mortality among hypoxic patients. For the immunomodulators dexamethasone (6mg daily), tocilizumab, and baricitinib, these results support their ongoing use in clinical practice for patients hospitalised with COVID-19 lung disease causing hypoxia.

The previously observed heterogeneity of dexamethasone treatment effect according to respiratory support status was also preserved over 6 months. The greatest proportional and absolute treatment benefit was seen in participants requiring invasive mechanical ventilation at randomisation with a smaller, but clinically important, benefit seen in those requiring oxygen only. Dexamethasone treatment remains unlikely to confer benefit, and may cause harm, in patients not requiring oxygen or when used at a higher dose among patients not receiving ventilatory support.

The addition of tocilizumab to corticosteroids or the addition of baricitinib to corticosteroids with or without IL-6 antagonists provided additional benefit. This supports treatment guidelines that suggest combining immunomodulatory therapy in participants at high risk of death from COVID-19.^6,7^ Since the mortality rate ratio benefit of these three treatments appears to be approximately independent (i.e. there was no evidence of an interaction by baseline use of other immunomodulators; appendix p88), for patients with COVID-19 and elevated CRP, their combined use could reduce mortality by about a third among those who are receiving oxygen (with or without non-invasive ventilation) and by about half among those requiring invasive mechanical ventilation. There is limited information on long-term outcomes from other trials of immunomodulators for severe COVID-19. The REMAP-CAP trial showed that IL-6 antagonists (tocilizumab and sarilumab) reduced 6-month mortality by about one quarter.^25^ The COV-BARRIER trial found that baricitinib reduced mortality at 2 months, but no longer term follow-up was reported.^26^

The availability of routine healthcare data allowed us to retrospectively identify patients who were severely immunocompromised at baseline and explore the effect of immunomodulatory treatments in this group for the first time. This did not indicate any hazard with immunomodulation using dexamethasone or baricitinib, but it did suggest a possible increase in all-cause mortality with tocilizumab, with no evidence that this was related to an increase in major non-COVID infection (appendix pp89-90). It is unclear why any hazard should be specific to IL-6 antagonism, although, if real, it could relate to the long duration of action of tocilizumab, which, unlike the other immunomodulators, is a monoclonal antibody.

There was some evidence of an increased risk of non-COVID infection among patients who received dexamethasone (but not tocilizumab or baricitinib), but the absolute excess was small (2%) and should be weighed against the mortality benefit in hypoxic patients. Corticosteroids exert an immunosuppressive effect by acting on many immune cells, including reducing macrophage and T-cell activation, and inhibiting pro-inflammatory cytokines.^27^ A large meta-analysis of 71 corticosteroid randomised controlled trials (for any indication) suggested an estimated 60% increased infection risk over a median of 21 days follow-up.^28^ In other acute diseases, such as pneumonia, septic shock, and traumatic brain injury, meta-analyses of randomised trials of short-term corticosteroids have not demonstrated an increased secondary infection risk, although these have had limited power to exclude small but potentially important effects.^29–31^ Our results suggest that for most patients in whom immunomodulation is indicated, the mortality benefit of treatment with dexamethasone is likely to offset the potential adverse effect on non-COVID infection.

Long-term use of tocilizumab and baricitinib are known to be associated with infections in randomised studies outside of COVID-19, predominantly in rheumatoid arthritis.^32– 34^ By contrast, we found no such evidence in the context of severe COVID-19. A meta-analysis of trials of IL-6 antagonists for COVID-19 found no evidence of an increased risk of secondary infections at short-term follow-up.^35^ Trials of JAK inhibitors in COVID-19, including RECOVERY, have also found no evidence of treatment-related infection risk at short-term follow-up.^3,36^ Prior to our results, data on infections after one month was too limited to draw conclusions (23 secondary infection events in a single trial).^37^ The lack of a substantial infection risk with either tocilizumab or baricitinib in RECOVERY and other COVID-19 randomised trials could be explained by the short duration of treatment, or represent the altered risk-benefit balance in the setting of acute severe illness.

Our results suggest that virus-directed monoclonal antibody therapies that are able to neutralise prevalent SARS-CoV-2 variants reduce all-cause mortality for hospitalised patients who are not mounting an adequate immune response (as evidenced by absent anti-viral-antibodies or high levels of viral antigen). There were no late hazards of neutralising monoclonal antibody treatment. The emergence of dominant viral variants with minimal susceptibility to casirivimab-imdevimab and sotrovimab has led to their withdrawal from treatment guidelines.^6^ However, the consistency of our results for two different monoclonal antibodies suggests that use of a monoclonal antibody for patients with a susceptible viral variant would be beneficial.

No other treatment evaluated in RECOVERY (aspirin, azithromycin, colchicine, convalescent plasma, dimethyl fumarate, empagliflozin, lopinavir-ritonavir, molnupiravir, and nirmatrelvir-ritonavir) showed clinical benefit at long-term follow-up, and both hydroxychloroquine and higher dose dexamethasone showed evidence of harm. In the REMAP-CAP trial there was some evidence that antiplatelet treatment reduced mortality and improved health-related quality of life at 6 months, but we did not find any evidence of a benefit of aspirin on 6-month mortality in RECOVERY.^25^ The moderate reduction in non-COVID infection we observed with azithromycin is in keeping with its antibiotic activity, but would not justify its routine use for COVID-19.

Strengths of this study included that it provided a large, randomised controlled comparison of a range of potential treatments. By the 6-month follow-up point, more than 98% of participants had either died or been discharged alive from hospital, giving an outcome for almost all participants from their initial hospital admission with COVID-19. Linkage to routinely collected UK healthcare data improved the completeness of follow-up and allowed ascertainment of additional baseline characteristics and trial outcomes. Limitations include the fact that RECOVERY was open label, so participants and local hospital staff were aware of the assigned treatment, which could potentially have affected clinical management or the recording of some trial outcomes, including diagnoses of infection. However, the primary and secondary trial outcomes are unambiguous and were ascertained without bias through linkage to routine health records in the large majority of patients. RECOVERY only studied patients who had been hospitalised with COVID-19 and, therefore, is not able to provide any evidence on the safety and efficacy of treatments in other patient groups, such as those with early infection. We also do not currently have data on the effects of these treatments on the development of long COVID.

In summary, among patients hospitalised with COVID-19, the early mortality benefits seen with dexamethasone 6mg daily, tocilizumab, and baricitinib, as well as monoclonal antibody treatments for those with a sensitive virus variant who are not mounting an adequate immune response, were sustained over 6 months.

## Data Availability

The protocol, consent form, statistical analysis plan, definition & derivation of clinical characteristics & outcomes, training materials, regulatory documents, and other relevant study materials are available online at www.recoverytrial.net. As described in the protocol, the Trial Steering Committee will facilitate the use of the study data and approval will not be unreasonably withheld. Deidentified participant data will be made available to bona fide researchers registered with an appropriate institution within 3 months of publication. However, the Steering Committee will need to be satisfied that any proposed publication is of high quality, honours the commitments made to the study participants in the consent documentation and ethical approvals, and is compliant with relevant legal and regulatory requirements (e.g. relating to data protection and privacy). The Steering Committee will have the right to review and comment on any draft manuscripts prior to publication. Data will be made available in line with the policy and procedures described at: https://www.ndph.ox.ac.uk/data-access. Those wishing to request access should complete the form available at this site.

## Writing Committee (on behalf of the RECOVERY Collaborative Group)

Peter W Horby*, Leon Peto*, Mark Campbell*, Rachel Wade*, Guilherme Pessoa-Amorim, Vanessa Tobert, Natalie Staplin, Jonathan R Emberson, Karl Wallendszus, William M Stevens, Andrew King, Rijo Kurien, Charles Crichton, Christopher Brightling, Benjamin Prudon, Christopher A Green, Sheri Thackoorcharan, Paul Hine, Tim Felton, Richard Stewart, Heinke Kunst, Andrew Ustianowski, J Kenneth Baillie, Maya H Buch, Saul N Faust, Thomas Jaki, Katie Jeffery, Edmund Juszczak, Marian Knight, Wei Shen Lim, Alan Montgomery, Aparna Mukherjee, Andrew Mumford, Kathryn Rowan, Guy Thwaites, Marion Mafham^†^, Richard Haynes^†^, Martin J Landray^†^

## Contributors

This manuscript was initially drafted by LP, MC, RH, PWH and MJL, further developed by the Writing Committee, and approved by all members of the trial steering committee. RW, JRE, PWH, MJL, RH and LP had access to the study data. RW and JRE accessed and verified the data. RW did the statistical analysis. PWH and MJL vouch for the data and analyses, and for the fidelity of this report to the study protocol and data analysis plan, and had final responsibility for the decision to submit for publication. PWH, NS, JRE, JKB, MHB, SNF, TJ, KJ, EJ, MK, WSL, AMo, AMuk, AMum, KR, GT, MM, RH, and MJL designed the trial and study protocol. LP, MC, GP-A, VT, KW, WMS, AK, RK, CC, CB, BP, CAG, ST, PH, TF, RS, HK, AU, the Data Linkage team at the RECOVERY Coordinating Centre, and the Health Records and Local Clinical Centre staff listed in the appendix collected the data. All authors contributed to data interpretation and critical review and revision of the manuscript.

## Data Monitoring Committee

P Sandercock, J Darbyshire, D DeMets, R Fowler, D Lalloo, M Munavvar (from January 2021), I Roberts (until December 2020), A Warris (from March 2021), J Wittes

## Declaration of interests

The authors have no conflict of interest or financial relationships relevant to the submitted work to disclose. No form of payment was given to anyone to produce the manuscript. The Nuffield Department of Population Health at the University of Oxford has a staff policy of not accepting honoraria or consultancy fees directly or indirectly from industry (see https://www.ndph.ox.ac.uk/files/about/ndph-independence-of-research-policy-jun-20.pdf).

## Acknowledgements

Above all, we would like to thank the thousands of patients who participated in this trial. We would also like to thank the many doctors, nurses, pharmacists, other allied health professionals, and research administrators at participating hospital organisations. Supported in the UK by staff at the National Institute for Health and Care Research (NIHR) Clinical Research Network, NHS DigiTrials, UK Health Security Agency, Department of Health & Social Care, the Intensive Care National Audit & Research Centre, UK Renal Registry, Public Health Scotland, National Records of Scotland, the Secure Anonymised Information Linkage at University of Swansea, and the NHS in England, Scotland, Wales and Northern Ireland. This work uses data provided by patients and collected by the NHS as part of their care and support.

The RECOVERY trial is supported by grants to the University of Oxford from UK Research and Innovation (UKRI) and NIHR (MC_PC_19056), the Wellcome Trust (Grant Ref: 222406/Z/20/Z) through the COVID-19 Therapeutics Accelerator, and by core funding provided by the NIHR Oxford Biomedical Research Centre, the Wellcome Trust, the Bill and Melinda Gates Foundation, the Foreign, Commonwealth and Development Office, Health Data Research UK, the Medical Research Council Population Health Research Unit, the NIHR Health Protection Unit in Emerging and Zoonotic Infections, and NIHR Clinical Trials Unit Support Funding. TJ is supported by a grant from UK Medical Research Council (MC_UU_00040/03). WSL is supported by core funding provided by NIHR Nottingham Biomedical Research Centre. Tocilizumab, casirivimab and imdevimab, sotrovimab, and empagliflozin were provided through support from Roche, Regeneron, GSK, and Boehringer Ingelheim, respectively. Colchicine for use in Indonesia was provided by Combiphar. The views expressed in this publication are those of the authors and not necessarily those of the NHS, the NIHR or the Department of Health and Social Care. For the purpose of Open Access, the author has applied a CC BY public copyright license to any Author Accepted Manuscript version arising from this submission.

## Notes

### Competing Interest Statement

The authors have declared no competing interest.

### Clinical Trial

ISRCTN50189673 and NCT04381936

### Author Declarations

UK: Cambridge East Research Ethics Committee (ref: 20/EE/0101) VietnamHospital for Tropical Diseases Ethics Committee Nepal: Ethical Review Board, Nepal Health Research Council (NHRC) Indonesia: Ethics Committee of the Faculty of Medicine, University of Indonesia Ghana: Ghana Health Service Ethics Review Committee South Africa: The University of the Witwatersrand, Human Research Ethics Committee India: ICMR Central Ethics Committee on Human Research (CECHR) The ethics committees/IRBs above gave ethical approval for this work. See appendix p31 for more details.

